# The impact of Test Positivity on Surveillance with Asymptomatic Carriers

**DOI:** 10.1101/2022.06.10.22276234

**Authors:** Mauro Gaspari

## Abstract

**Objectives:** Recent studies show that Test Positivity Rate (TPR) gain a better correlation than incidence with the number of hospitalized patients in COVID-19 pandemic. Nevertheless, epidemiologist remain sceptical concerning the widespread use of this metric for surveillance, and indicators based on known cases like incidence are still preferred despite the large number of asymptomatic carriers which remain unknown. Our aim is to compare TPR and incidence, to determine which of the two has the best characteristics to predict the trend of hospitalized patients in COVID-19 pandemic.

**Methods:** We perform a retrospective study considering 60 cases, using global and local data from Italy in different waves of the pandemic, in order to detect peaks in TPR time series, and peaks in incidence, finding which of the two has the best ability to anticipate peaks in patients admitted in hospitals.

**Results:** On average the best TPR based approach anticipates incidence of about 4.6 days (95% CI 2.8, 6.4), more precisely the average distance between TPR peaks and hospitalized peaks is 17.6 days (95% CI 15.0, 20.4) with respect to 13.0 days (95% CI 10.4, 15.8) obtained for incidence. Moreover, the average difference between TPR and incidence increases to more than 6 days in the Delta outbreak during Summer 2021, where presumably the percentage of asymptomatic carriers was larger.

**Conclusions:** We conclude that TPR should be used as primary indicator to enable early intervention and for planning hospitals admissions in infectious diseases with asymptomatic carriers.

## 1 Introduction

Test positivity rate (TPR), e.g, the percentage of positive tests over total tests, is one of the metrics used for public health surveillance in infectious diseases. Typical applications include estimating the prevalence of diseases in the population, see [1] for malaria disease and [2] for COVID-19, or establish levels of community transmission from sentinel sites in the COVID-19 pandemic [3]. Over the past two years, thanks to the large amount of data collected in the COVID-19 pandemic, new application domains where explored to guide epidemiologic policy-making [4, 5, 6] assessing epidemic dispersal mediated by asymptomatic carriers. Recent studies have highlighted a correlation with the number of patients admitted in hospitals [7, 8], which increases with respect to other indicators like incidence [9] or the daily number of positive cases [10]. This kind of correlation, was exploited to forecast two weeks in advance variations on the number of patients admitted in hospitals on the basis of TPR variations [11], or to define a severity detection rate, to predict ICU admissions [12].

Despite these promising results, the large percentage of asymptomatic carriers in COVID-19 [13, 14], and the risk of hampering control efforts [15], epidemiologist remain sceptics concerning the widespread use of TPR for surveillance, and other indicators based on known cases like incidence (7-day incidence rate per 100,000) are in general preferred. One of the motivations of this choice is that the calculation of TPR is more critical and there is still little agreement on the method to be used.

A general issue is that tests are not usually classified in the statistics, despite they belong to different categories, and only the total number of administered tests is reported daily. Indeed, conducted tests include both diagnostic tests administered with the goal of discovering new cases; and control tests, addressed to infected individuals, to monitor the course of the disease or to check the healing. While the positive percentage of the former can be used for surveillance modelling the progress of the pandemic, the positive control tests should be used for different purposes, for example to evaluate the length of quarantine [16]. Due to this lack of information, test positivity is usually computed as the ratio between new positive cases and the total number of tests done [17], being only an approximation of the actual positivity rate.

Another open issue is whether or not antigen tests should be part of the calculation. For example, CDC (US Centers for Disease Control and prevention) computes test positivity as the percentage of all SARS-CoV-2 Nucleic Acid Amplification Tests (NAAT) conducted that are positive, while they recommend to collect antigen tests as separate data[17]. On the contrary, several available statistics use both of them as denominator, such as those presented in the Coronavirus Testing web site [18]. Moreover, when antigen tests are used in the TPR calculation an additional problem arises, health care guidelines may recommend that positive antigen tests should be confirmed by NAAT tests because the latest have a better accuracy. These repeated diagnostic tests done for the same positive individuals should also be removed by the denominator [8].

Furthermore, there are several problems related to data collection in different regions or countries. For example, in most of the Italian regions the number of test reported on Monday is lower than the those reported in other days of the weak and includes a lower ratio of antigen tests. From this it follows that, the computed TPR is in general higher on Monday with respect to the other days of the week. In other words in Italy there is a form of weekly seasonality in TPR time series, and similar problems also arise in other countries.

Finally, a negative correlation between TPR and the number of administered tests was evidenced in some studies [19, 4, 12]. Basically, when the number of conducted tests increases, test positivity tends to decrease, and thus some variations of TPR may be linked to an increased volume of testing. This phenomenon is more evident when the number of administered tests per million inhabitants is low, and tends to mitigate when a large number of tests are conducted. For example in fall 2021 when green pass for workers became mandatory in Italy, and the number of performed tests almost doubled in a few days, a significant variation was not observed in the TPR time series. To deal with this issue an adjusted TPR calculation method was proposed in [19] to be used in countries where capacity of testing is limited.

Our first goal is to clarify TPR calculation issues to identify which method behaves better for surveillance purposes. Namely, which method has the best performance in predicting the trend of hospitalized patients. Then, our aim is to compare TPR and incidence to determine which of the two has the best characteristics for the same purpose. The metrics that we use for this comparison is based on the intuition that an optimal indicator for planning hospitals admissions should be able to track the progress of infections. More precisely, the number of infections that occur day by day in a pandemic: if the number of infections increases an optimal indicator should also increase, while vice-versa, if the number of infections decreases, an optimal indicator should decrease. This aspect cannot be modelled by Rt (the reproduction number) only, because prevalence levels should be considered to estimate the volume of infections. Indeed Rt only represents the number of secondary infections generated from a case day by day, and not the total number of infections.

The peaks of an indicator that models the trend of infections should precede the peaks in hospitalized patients time series. Indeed hospital admission always occurs several day after infection, including incubation period, symptom onset and patient testing, about 15 days for COVID-19 [11], possibly more. The same consideration holds for admission in intensive care units. Thus, we consider the distance in days from the peaks of hospitalized time series as a metric to measure the predictive capacity of the analysed indicators.

In this paper, we compare incidence with different TPR calculation methods: standard rolling averages, the methods defined in [8, 11], the adjusted TPR proposed in [19], and a new proposal based on a two level approach. We consider 60 different cases using global and local data from Italy in four waves of the pandemic, starting from the second wave where data on antigen tests were made available for some region (Fall 2020), and the successive Alpha, Delta and Omicron waves.

In summary, we aim to answer some key questions concerning COVID-19 and infectious diseases in general: is incidence the most appropriate indicator to track infections when many asymptomatic carriers are present? would TPR have similar or better performance in the same context? which TPR calculation method should be used? should antigen tests be used in the TPR calculation?

## 2 Methods

Unfortunately, since tests are not classified in Italian official data (only NAAT and antigen tests are distinguished [25]), in this study we have to approximate the TPR as the ratio between new positive cases and the total number of tests done, this solution is a common approach used in many countries [18], also recommended in [17].

Starting from this basic calculation method, we compare 7 different versions of TPR, two of them are based on NAAT nasopharyngeal swab only, and the other exploit antigen tests. The first 6 versions are obtained computing the rolling average of the last 7 days, a common practice to address anomalies in data collection [18], of the following ratios:

N1: New Positive cases / NAAT tests only.

N2: New positive cases detected with NAAT tests only / NAAT Tests only.

A1: New Positive cases / NAAT + Antigen tests.

A2: New Positive cases / NAAT + Antigen tests - Estimated Repeated Tests.

A3: New Positive cases / NAAT + Antigen tests - Number of healed patients.

A4: New Positive cases / NAAT + Antigen tests * (growth rate of cases / growth rate of tests).

Version A1 is the usual 7 days rolling average based on all the tests done, while versions A2 and A3 are attempts to improve its accuracy removing from the denominator tests that are not devoted to the diagnosis of new cases, as follows:

A2 It removes an estimation of repeated diagnostic tests (NAAT tests conducted to confirm positive Antigen tests) computed using the approach presented in [11].

A3 It removes the number of healed people, assuming that at least one test was administered to each of them [11]. This data is usually reported daily in most of the countries.

Version A4 is the adjusted TPR presented in [19] which deals with the negative correlation between TPR and the number of administered tests. See Appendix A for a detailed description of the above TPR calculation methods.

In addition, we also evaluate a new version of TPR (A5) based on a two level approach which aims to model with better accuracy the progress of infections, generating more stable time series.

### 2.1 A two level TPR calculation method

A criticism to methods based on standard 7 days rolling average, which compute the TPR of a given day as the average of the preceding 7 days, is that they mainly deal with data collection issues, without considering modelling and epidemiological issues. Indeed, from a knowledge modelling perspective estimating TPR of a given day as the average of preceding and following days, will be more appropriate and correct. Moreover, 7 days are necessary to deal with week seasonality issues, but significant variations in the progress of infections could be captured with better accuracy looking at less than 7 days. For example, the length in days of the incubation period, which represents the minimal number of days after which changes can be observed.

To get through these issues, we have devised a two level approach for calculating the TPR: the first level addresses modelling issues, and in the second epidemiological issues. Let *t*_1_,*t*_2_,… *t*_*n*_ be the daily TPR time series, the TPR at time *i*, when *i >* 3 and *i < n −* 3 can be modelled computing the trend as follows:

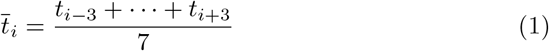

Namely, the TPR in a given day is modelled computing the average value of the days preceding and following it. This formula cannot be computed for the last three days, but it can be approximated computing averages on the available days only, as follows: 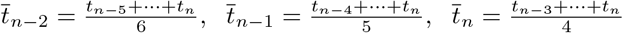 where *t*_*n*_ be the daily TPR of the last day.

However, this simple approach, which assumes that the average TPR value will not have significant changes in the remaining days, does not work well due to the seasonality of the daily TPR time series. Figure 1 (a) illustrates this problem considering the Emilia Romagna region: the highest values of daily TPR occur on Monday while on Tuesday the daily TPR is well below average. Thus, if the last day is Monday, the TPR will be overestimated, and if it is Tuesday probably underestimated.

**Figure 1:**
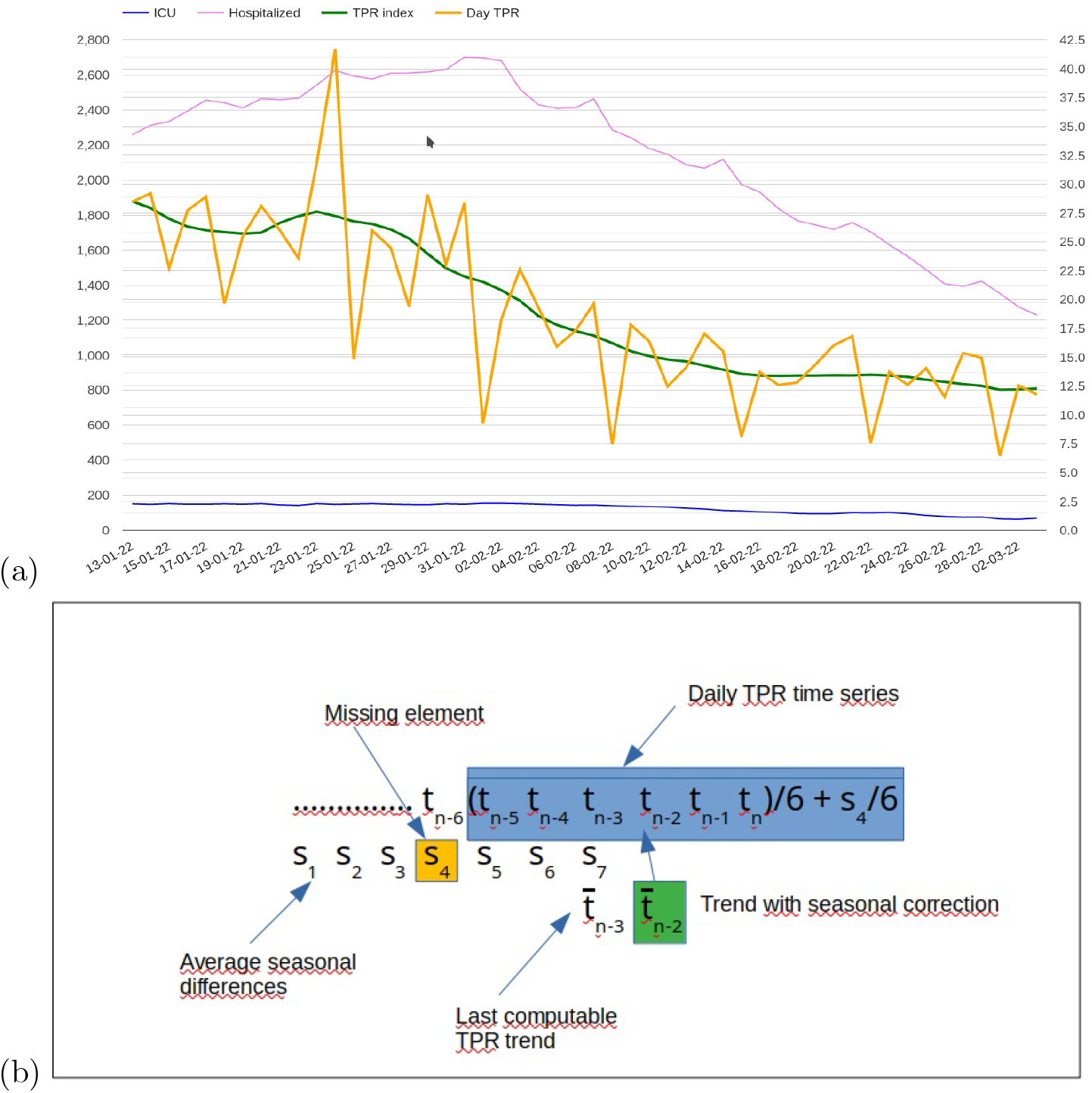
(a) Daily TPR index seasonality in Emilia Romagna region January/February 2022; (b) Computing the TPR trend of the last 3 days adding seasonal corrections: 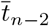 example.

To solve this problem we compute the difference between the TPR trend (computed with formula 1) and daily TPR for each day in the last k weeks. Let (*s*_1_,*s*_2_,*s*_3_,*s*_4_,*s*_5_,*s*_6_,*s*_7_), be the list of average values of these differences for each day in the week (where *s*_7_ is the mean difference associated to the last element of the trend time series 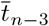), the TPR of the last 3 days is obtained adding to the previous formulas the weighted differences of the days that are missing (e.g. not used for computing the formula 1, see Figure 1 (b)): 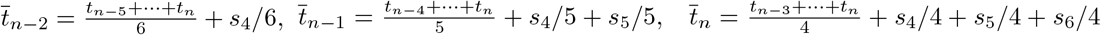. The experiments we did show that this approach avoids the effects of seasonality estimating TPR in the last 3 days. More precisely, analysing time series history (more than 16000 cases), setting k=4 weeks: 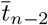 improves the naive approach on average of 0.0004, 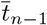 of 0.0022, and 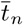 of 0.0032. Increasing or reducing k worsens these results. It should be noted that this issue does not have an influence on the peaks of the TPR time series, thus it does not interfere with the metric used in the study.

Starting from the trend time series computed as above, we introduced a second level to compute the final TPR value considering epidemiological issues. The TPR value is obtained as the average of the last *µ* days of the trend time series, where *µ* is the incubation period.

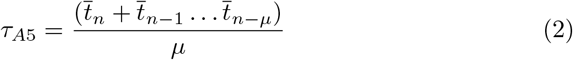

Where *µ* = 5 initially [21], and *µ* = 3 in the Omicron outbreak [22, 23].

We will show how this approach effectively improves the predictive properties and the regularity of the TPR time series.

### 2.2 Data Collection

The data used for this study were made available by the Italian Department of Civil Protection [25] for all the Italian regions, for all the course of the pandemic. This site contains all the relevant time series needed for our analysis, in details: new positive cases; NAAT tests; antigen tests; recovered, patients admitted in hospital and in ICU. However, the structure of the dataset was changing overtime, and data fundamental for our analysis were not always available and/or reliable, therefore for each wave of the pandemic some of the regions were discarded due to lack of data or known reliability issues.

The first wave of the pandemic starting from February 2020 was not considered in the study because antigen tests were not used in Italy. In the second wave (from the 1st of October 2020 to the 10th of January 2021) the data on antigen tests were not available in the official site, however they have been made available for 5 regions: Toscana, Piemonte, Friuli Venezia Giulia, Veneto and the Provincia Autonoma of Bolzano [8]. Since, some of the data collected for Veneto, Friuli and Bolzano are uncertain (see [8] for data collection details), only Toscana and Piemonte were considered in this study, and all the other regions were excluded from the analysis.

From the 15th of January 2021 onwards, data on antigen tests have been made available for all the regions of Italy. In spite of this, in the successive Alpha wave (from the 10th February 2021 to the 1st of May 2021) which started right after (February 2021), some regions were still excluded because hospitalized patients had started to grow before the beginning of the observed period, where the data on tests were still unreliable. The excluded regions are: Abruzzo, Umbria, Molise, and Basilicata.

As regard the Delta wave (from the 1st of July 2021 to the 10th of October 2021), all the regions of Italy were considered except Lazio, where data on tests were damned due to a hacker attack.

Successively, in the last Omicron outbreak (from the 24th of December 2021 to the 18th of February 2022): Sardegna was excluded since the TPR peak was at the end of the observed period (the 16th of February); Valle d’Aosta was excluded due to an error in the reported data concerning positive cases detected with NAAT test only; Provincia of Bolzano was excluded because a large amount of antigen tests were not reported in this period.

Finally, global data of Italy in the last 3 waves were also considered as further case studies, obtaining a total of 60 different cases: 2 regions in the second wave, 17 regions and the whole Italy in the third wave, 20 regions and the whole Italy in the Delta wave, and 18 regions and Italy in the Omicron wave.

Although all the data come from a single country, Italy, we believe that the collected sample is sufficiently general and heterogeneous in order to draw valid conclusions. Indeed, Italian, regions ranges from little territories with less then 500 thousand inhabitants to larger regions having over 10 million inhabitants. Moreover, Italian regions have their own health departments and different organizations, and, as a consequence, heterogeneous data collection policies for the administration of diagnostic tests.

### 2.3 Data Analysis

For all the analysed cases, we compute the number of days that occur between peaks of the above indicators and peaks of hospitalized people considering both patients admitted in non critical areas and in intensive care units. We analyse the generated samples computing average values and standard deviation, also considering differences between indicators, and the effects due to outliers. We use the Shapiro-Wilk test to check normality [29], and, when the test fails, we use a simple non-parametric Bootstrap method [20] with Monte-Carlo simulation performing 5000 iterations, to compute confidence intervals for the obtained averages values.

Moreover, we use sample entropy [27] as a measure of the regularity of these indicators [28]. This is a measure of the probability that two segments of the analysed time series within a given period minimize discontinuities. Smaller values of sample entropy indicates a greater probability that a set of TPR values will be followed by similar values, while a larger value indicates major irregularities. We assume 2 as embedded dimension and the Chebyshev distance as a metric.

## 3 Results and Discussion

The results we obtained considering all the selected 60 cases are summarised in Table 1. An initial analysis of these results allows us to draw preliminary conclusions about the different versions of TPR, specifically: N2, which uses positives cases determined with NAAT tests only as a numerator, outperforms N1 which uses all the new positives; A1, e.g. the standard 7 days rolling average including both NAAT and antigen tests, outperforms versions A2, A3 and the adjusted TPR A4. Basically, all the attempt to improve TPR accuracy removing tests from the denominator, or considering changes in the number of administered tests seem to fail. For example, removing tests used to check the healing, apparently provides a better approximation during the growth phase of the curve, but it tends to create artificial peaks, before the phase of descent, when the number of recovered patients is high.

**Table 1:** This table summarises the average distances between peaks of the studied indicators with respect to patients admitted in non-critical areas and ICUs, considering all the 60 cases. We present average values, standard deviation and sample entropy for each indicator.

| Indicator | Hospitalized |  | Sample Entropy | ICU |  |
| --- | --- | --- | --- | --- | --- |
|  | *Avg Dist.* | *Std Dev.* |  | *Avg Dist.* | *Std Dev.* |
| **Incidence** | 13.067 | 10.734 | 0.305 | 10.417 | 13.631 |
| **TPR N1** | 11.7 | 12.17 | 0.34 | 9.05 | 14.835 |
| **TPR N2** | 13.833 | 11.649 | 0.421 | 11.183 | 12.614 |
| **TPR A1** | 16.083 | 11.279 | 0.324 | 13.433 | 15.692 |
| **TPR A2** | 16.033 | 11.309 | 0.327 | 13.383 | 15.662 |
| **TPR A3** | 13.65 | 11.828 | 0.349 | 11.0 | 16.813 |
| **TPR A4** | 16.033 | 11.237 | 0.334 | 13.383 | 15.192 |
| **TPR A5** | 17.633 | 10.66 | 0.225 | 14.983 | 14.871 |

Last but not least, the new version of TPR (A5) outperforms all the other indicators including incidence and has a better regularity, considering both standard deviation and sample entropy. Thanks to these properties, a decrease observed after a growth phase has more chance of identifying a peak, rather than an anomalous effect. Figure 2 presents a comparison between A5 and the naive 7 days rolling average (A1) illustrating the benefits of having a better sample entropy.

**Figure 2:**
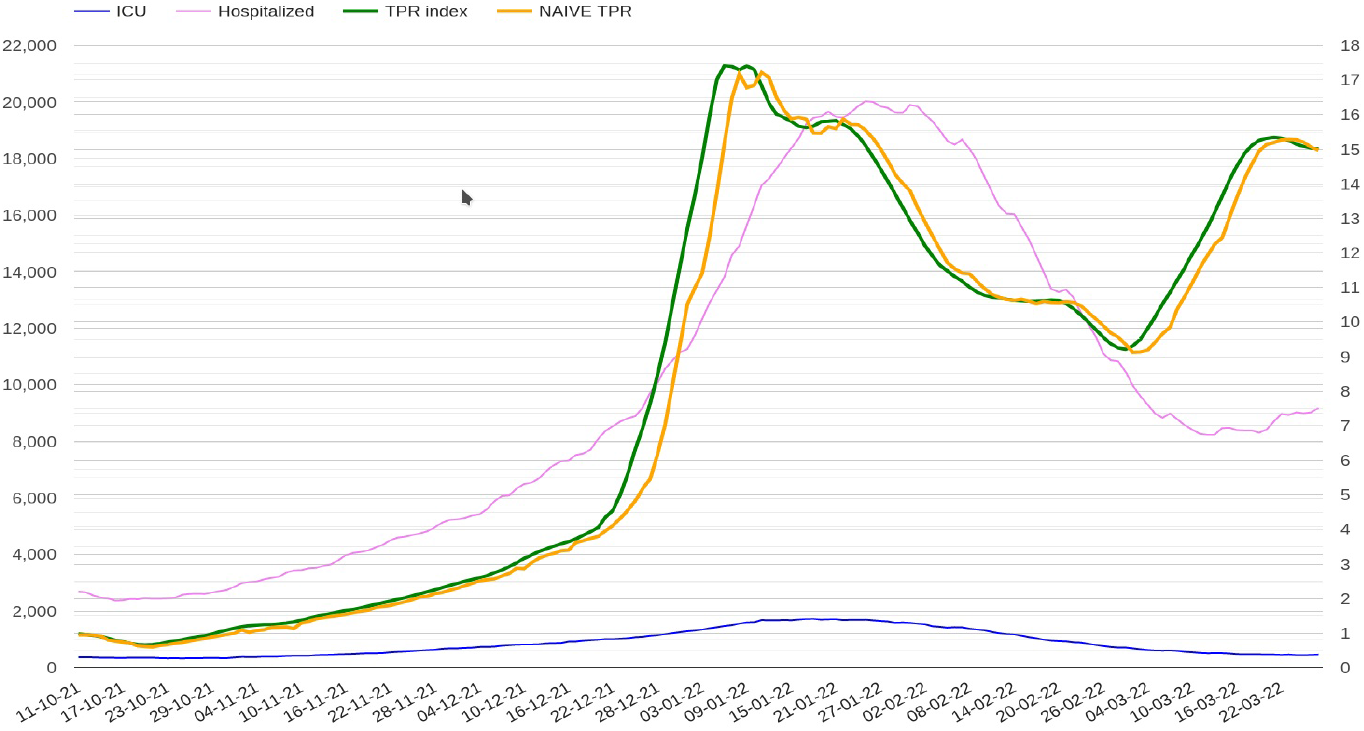
The best TPR (A5) computed for Italy compared with naive 7 days rolling average (A1) in the Omicron phase.

These considerations for TPR versions N1, A1, and A5 hold in all the analysed waves, thus we focus the subsequent discussion on these indicators only comparing them with incidence. Similar results hold for both patients admitted in non critical areas and in intensive care units. However, considering ICUs it is clear that the results do not allow to reach meaningful conclusions, because standard deviation is quite high, thus a more deep analysis is needed for ICUs.

If we consider patients admitted in non critical areas instead, it appears that TPR (A5) has a better predictive capacity than incidence when asymptomatic carriers are present, providing a preliminary answer to one of the key questions we asked at the beginning of the paper. Indeed, the average distance between TPR peaks and hospitalized peaks is 17.6 days with respect to 13.0 days only, obtained for incidence. In practice the best TPR based approach anticipates incidence of about 4.6 days. These preliminary observations are certainly strengthened by the fact that we obtained similar results computing average values of TPR and incidence in all the considered waves, that is, TPR outperforms incidence in all the analysed waves, and the results have similar proportions. However, more considerations are needed to draw definitive conclusions and compute confidence intervals.

Let Δ_*T P R*_ and Δ_*I*_ be respectively the sets of distances between TPR (version A5) and incidence peaks with respect to peaks of hospitalized time series, and Δ_*T P R−I*_ be the set of differences between Δ_*T P R*_ and Δ_*I*_ case by case. Figure 3 (a,b,c) shows how the elements of these sets are distributed. Although their distributions are not far from Gaussian, the Shapiro-Wilk test of normality [29] returns p-values less than .05, with sufficient evidence that data do not come from normal distributions. More precisely: Δ_*T P R*_ stat: 0.958, pvalue: 0.041; Δ_*I*_ stat: 0.942, pvalue: 0.007; Δ_*T P R−I*_ stat: 0.928, pvalue: 0.001). However, the analysis of single cases has shown that extreme cases are always outlier, thus the considered sets do not belong from heavy-tailed distributions. In light of these concerns, we use a non-parametric Bootstrap method with Monte-Carlo simulation to estimate confidence interval of averages of the analysed distribution (Figures 3 (d,e,f). The experiments we did show that 5000 iterations are enough to converge generating stable values, the resulting confidence intervals are: Δ_*T P R*_: 95% CI 15.0, 20.4; Δ_*I*_ : 95% CI 10.4, 15.8; Δ_*T P R−I*_ : 95% CI 2.8, 6.4.

**Figure 3:**
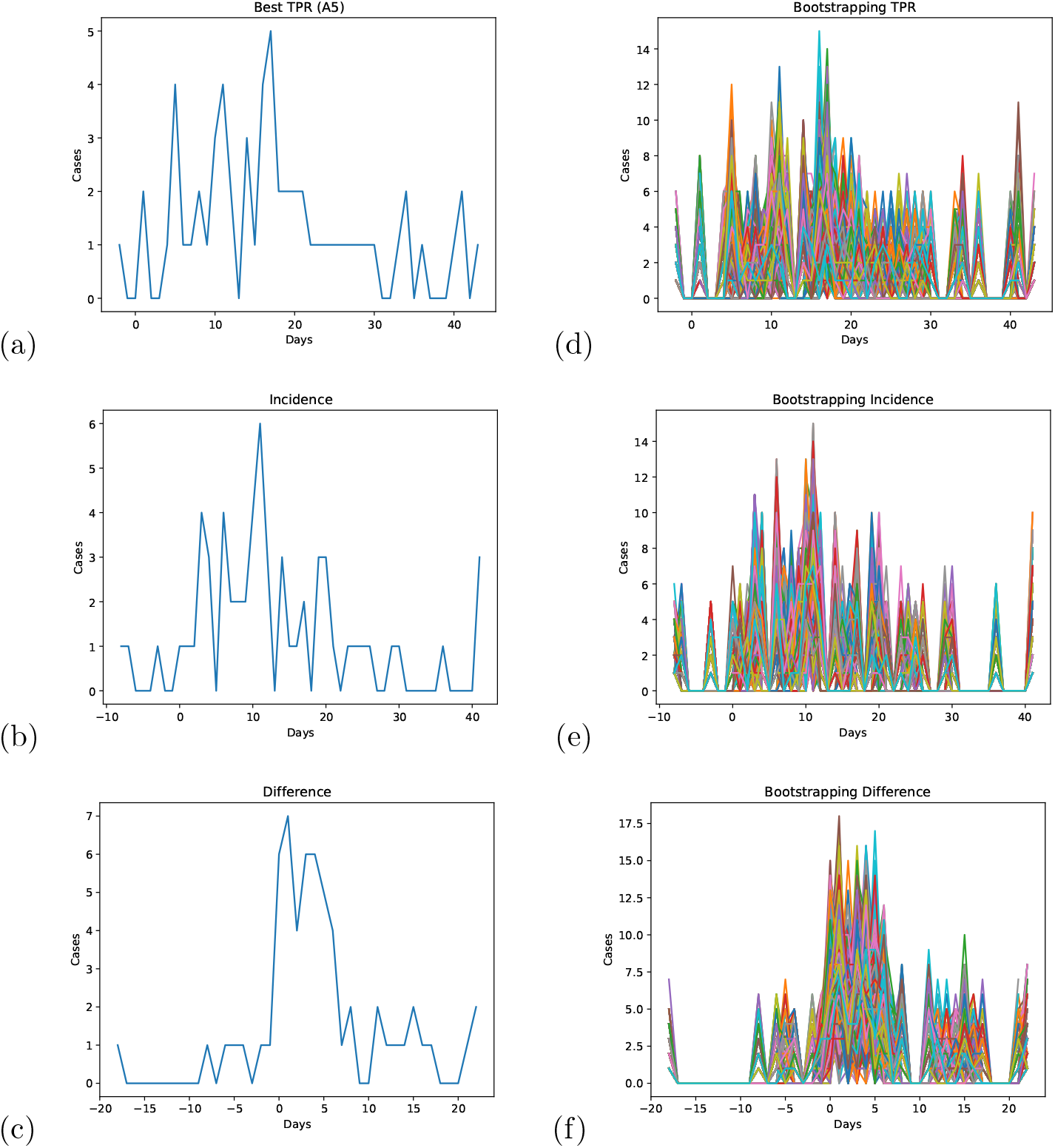
The figure shows how the values in the sets Δ_*T P R*_, awg: 17.6, sd: 10.7 (a), Δ_*I*_, awg: 13.1, sd: 10.7 (b), and Δ_*T P R−I*_, awg: 4.6, sd: 7.1 (c), are distributed, and the results obtained boostrapping them with 5000 iterations (d,e,f).

Given that confidence intervals estimated for TPR and incidence overlap, we analyse all single cases where incidence anticipates TPR. As Figure 3 (c) shows, incidence anticipates TPR of more than 2 days in 5 out of 60 cases only. Nonetheless, in all these cases which include: Lazio (18 days), Campania (4) and Sardegna (5) in the 3rd wave, Lombardia (8) in the Delta wave, and Puglia in the Omicron wave, the trends of TPR and incidence remain similar, both of them reach the top of a plateau about at the same time, and the delay of TPR peaks are caused by small variations in the plateau.

Analysing instead the relationship of TPR and incidence with hospitalized patients, TPR peaks follows peaks of hospitalized patients in one case only (Friuli in the Delta wave), while this happens three times for incidence (Friuli, Valle d’Aosta, and Marche in the Delta wave).

In summary, although there are a few cases where incidence anticipates TPR, their average 95% confidence intervals are almost disjoint, and it never happens in practice that incidence really outperforms TPR, with a possible negative impact on surveillance. This means that techniques that exploit TPR for estimating two weeks in advance variations in hospital admissions, like those presented in [11], will work anyway for the detected outliers. We can therefore conclude that for surveillance purposes TPR outperforms incidence in COVID-19. The insight of this research is that metrics based on known cases are not able to model with sufficient accuracy the progress of infections in infectious diseases with asymptomatic carriers, while TPR also accounting for unknown cases, and thus modelling under-ascertainment [26] does it.

Concerning intensive care units an in-depth analysis of the obtained results shows that they were negatively influenced by low values found for the Omicron variant (19 cases), as presented in Table 2 (a). Indeed, if we exclude the Omicron wave, and we compute the average distances between peaks for the remaining 41 cases, the results get closer to those obtained for patients in non-critical areas (see Table 2 (b)). Indeed, the best version of the TPR anticipates incidence of about 4.2 days, in details the average distance between TPR and ICU peaks is about 18.3 days and 14 days only for incidence. On the contrary, for hospitalized in non-critical areas in the Omicron outbreak average values are conform to those obtained in the other waves. Anyway, the values of standard deviation for ICUs remains high.

**Table 2:** TPR and Incidence average peaks distances with respect to patients admitted in non critical areas and ICUs considering different waves and seasons. For each indicator we present average values, standard deviation and sample entropy.

**(a) Omicron wave (19 cases)**
| Indicator | Hospitalized |  | Sample Entropy | ICU |  |
| --- | --- | --- | --- | --- | --- |
|  | *Avg Dist.* | *Std Dev.* |  | *Avg Dist.* | *Std Dev.* |
| **Incidence** | 12.895 | 8.687 | 0.332 | 2.579 | 13.019 |
| **TPR N2** | 15.263 | 12.9 | 0.524 | 4.947 | 15.582 |
| **TPR A1** | 15.474 | 10.475 | 0.237 | 5.158 | 14.612 |
| **TPR A5** | 18.158 | 10.137 | 0.2 | 7.842 | 14.091 |

**(b) All the cases excluding Omicron wave (41 cases)**
| Indicator | Hospitalized |  | Sample Entropy | ICU |  |
| --- | --- | --- | --- | --- | --- |
|  | *Avg Dist.* | *Std Dev.* |  | *Avg Dist.* | *Std Dev.* |
| **Incidence** | 13.146 | 11.56 | 0.292 | 14.049 | 12.317 |
| **TPR N2** | 13.171 | 10.959 | 0.373 | 14.073 | 9.694 |
| **TPR A1** | 16.366 | 11.622 | 0.365 | 17.268 | 14.662 |
| **TPR A5** | 17.39 | 10.885 | 0.236 | 18.293 | 14.037 |

**(c) Delta wave, summer 2021 (21 cases)**
| Indicator | Hospitalized |  | Sample Entropy | ICU |  |
| --- | --- | --- | --- | --- | --- |
|  | *Avg Dist.* | *Std Dev.* |  | *Avg Dist.* | *Std Dev.* |
| **Incidence** | 14.762 | 14.573 | 0.32 | 16.952 | 15.37 |
| **TPR N2** | 14.19 | 13.971 | 0.372 | 16.381 | 11.902 |
| **TPR A1** | 20.048 | 13.899 | 0.359 | 22.238 | 17.765 |
| **TPR A5** | 21.143 | 12.631 | 0.252 | 23.333 | 16.915 |

**(d) Cold seasons: non-critical areas (39 cases); ICUs excluding the Omicron outbreak (20 cases).**
| Indicator | Hospitalized |  | Sample Entropy | ICU |  |
| --- | --- | --- | --- | --- | --- |
|  | *Avg Dist.* | *Std Dev.* |  | *Avg Dist.* | *Std Dev.* |
| **Incidence** | 12.154 | 7.781 | 0.296 | 11.0 | 6.693 |
| **TPR N2** | 13.641 | 10.177 | 0.447 | 11.65 | 5.695 |
| **TPR A1** | 13.949 | 8.869 | 0.306 | 12.05 | 7.493 |
| **TPR A5** | 15.744 | 8.871 | 0.21 | 13.0 | 6.986 |

A plausible explanation of the low predictive values obtained in the Omicron outbreak for both TPR and incidence with respect to ICU, is that this effect is due to the concatenation of Delta and Omicron variants. A reasonable hypothesis is that the observed peaks for ICU, which occurred in the first half of January, mostly depend from Delta peaks presumably occurring at the end of December 2021. Subsequently, when Omicron has become dominant, there was a significant reduction of critical cases confirmed by an evident decrease of the ratio between critical cases admitted in ICU and patients in non-critical areas, which globally in Italy dropped by half from the end of December (about 13%) to the first half of January (about 6%).

Another aspect which may give further guidance on the relationship of the studied indicators with asymptomatic carriers, is whether significant changes can be observed in different seasons of the year, for example during the summer when the number of asymptomatic carriers is presumably higher. Table 2 (c) presents the results of the analysis for the Delta outbreak at the beginning of July 2021. If we compare this results with those obtained in cold seasons presented in Table 2 (d), we can observe that the average time lags between TPR (A5) peaks and hospitalized time series increases from 15.7 to 21.1 days. This effect depends on at least two factors: 1) the course of the diseases which is presumably longer during the summer, 2) the percentage of asymptomatic which increases with hot weather. The first factor is supported by the fact that all the indicators increase, the second factor most likely depends on loss of accuracy of incidence in modelling the progress of infection due to a large proportion of asymptomatic carriers. It is worth noticing that during the winter (Table 2 (d) excluding the omicron outbreak) the results obtained for ICUs are more conform to those obtained in other studies like [11, 12], and standard deviation has acceptable values.

## 4 Conclusions

The results we have discussed, allows us to answer with sufficiently convincing arguments the questions asked at the beginning of this study. First, there is an evidence that incidence is not the best indicator for surveillance in infectious diseases when a considerable percentage of asymptomatic carriers is present, as for the COVID-19 pandemic, test positivity should be used instead. This result holds for patients admitted in non critical areas, while more investigations are needed for ICUs. Second, further support is given to the hypothesis that antigen tests should be used in TPR calculation. The performance of the best PCR based TPR calculation method are similar to those of incidence: the average is slightly higher, but standard deviation gets worse. In other words TPR outperforms incidence only if antigen tests are considered in the calculation. Key practical implications of this research are that: data collection procedures should be improved to make TPR calculation as accurate as possible; TPR based approaches to compute epidemiological parameters, like Rt, should be investigated more deeply.

## Data Availability

All data produced in the present study are available upon reasonable request to the authors

## Data availability

The data used in this study were provided by the Italian Civil Protection Department, and are available here: https://github.com/pcm-dpc. We also provide upon reasonable request the used dataset ()a csv file) and the computed time series using a google graph html format, which can be visualized using a simple Web browser.

## Ethics approval

This study does not involve humans or animals, no ethical approval is required.

## Consent to participate and for publication

This study does not involve humans or animals, no consent to participate or publication is required.

## A TPR calculation methods

Let *dayP* and *dayN* be respectively the new positive cases and cases detected with NAAT tests only for each day; let *dayT* and *dayA* be respectively the number of NAAT tests and the number of Antigen tests done for each day; let *dayR* and *Pr* be respectively the number of healed patients and an estimation of repeated tests, and let the notation 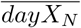 indicate N days average of *dayX*, we can define the above TPR versions as follows:

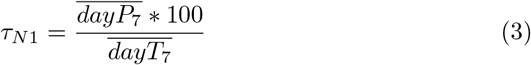

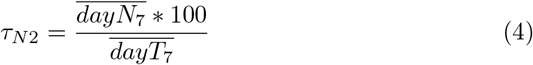

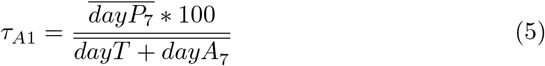

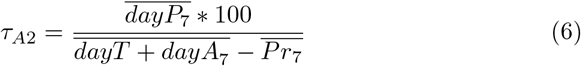

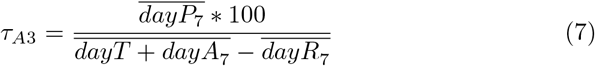

We brefly recall the method presented in [11] to estimate the average number of repeated tests *Pr*. Let 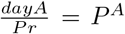 be the antigen positivity rate, and 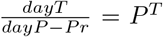 the NAAT positivity rate, given that in general *P*^*T*^ = *ψ P*^*A*^ _*dayP –Pr*_ where *ψ >* 1 holds, (namely the positivity rate for antigen tests is lower, see for example the results presented here [24]), if we set *ψ* = 2 which is a reasonable approximation observing the data that are available for Italy, we obtain:

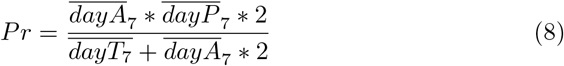

Finally, the adjusted TPR [19] is defined multiplying observed TPR, version *A*1, with a factor which express the growth rates of cases and tests as follows:

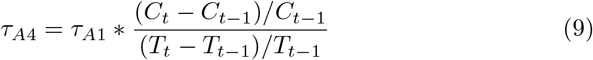

where *C*_*t*_ and *T*_*t*_ are the cumulative number of cases and tests at time *t* respectively.

